# Comparison of Foundation and Supervised Learning-Based Models for Detection of Referable Glaucoma from Fundus Photographs

**DOI:** 10.1101/2025.08.21.25334170

**Authors:** Kyle Bolo, Tran Huy Nguyen, Sreenidhi Iyengar, Zhiwei Li, Van Nguyen, Brandon J. Wong, Jiun L. Do, Jose-Luis Ambite, Carl Kesselman, Lauren P. Daskivich, Benjamin Y. Xu

## Abstract

**Purpose:** To compare the performance of a foundation model and a supervised learning-based model for detecting referable glaucoma from fundus photographs.

**Design:** Evaluation of diagnostic technology.

**Participants:** 6,116 participants from the Los Angeles County Department of Health Services Teleretinal Screening Program.

**Methods:** Fundus photographs were labeled for referable glaucoma (cup-to-disc ratio ≥ 0.6) by certified optometrists. Four deep learning models were trained on cropped and uncropped images (Training N = 8,996; Validation N = 3,002) using two architectures: a vision transformer with self-supervised pretraining on fundus photographs (RETFound) and a convolutional neural network (VGG-19). Models were evaluated on a held-out test set (N = 1,000) labeled by glaucoma specialists and an external test set (N = 300) from University of Southern California clinics. Performance was assessed while varying training set size and stratifying by demographic factors. xRAI was used for saliency mapping.

**Main Outcome Measures:** Area under the receiver operating characteristic curve (AUC-ROC) and threshold-specific metrics.

**Results:** The cropped image VGG-19 model achieved the highest AUC-ROC (0.924 [0.907-0.940]), which was comparable (*p* = 0.07) to the cropped image RETFound model (0.911 [0.892-0.930]), which achieved the highest Youden-optimal performance (sensitivity 82.6%, specificity 88.2%) and F1 score (0.801). Cropped image models outperformed their uncropped counterparts within each architecture (*p* < 0.001 for AUC-ROC comparisons). RETFound models had a performance advantage when trained on smaller datasets (N < 2000 images), and the uncropped image RETFound model performed best on external data (*p* < 0.001 for AUC-ROC comparisons). The cropped image RETFound model performed consistently across ethnic groups (*p* = 0.20), while the others did not (*p* < 0.04); performance did not vary by age or gender. Saliency maps for both architectures consistently included the optic nerve.

**Conclusion:** While both RETFound and VGG-19 models performed well for classification of referable glaucoma, foundation models may be preferable when training data is limited and when domain shift is expected. Training models using images cropped to the region of the optic nerve improves performance regardless of architecture but may reduce model generalizability.

## Introduction

Glaucoma is the leading cause of irreversible blindness worldwide.^1^ Its prevalence is rapidly rising due to population aging with a projected increase in the United States from 2.7 million cases in 2010 to 6.3 million in 2050.^2^ Within the current paradigms of eyecare delivery, the detection of asymptomatic glaucoma primarily relies on opportunistic detection during routine eye examinations. This practice pattern is estimated to result in the underdiagnosis of at least half of glaucoma cases.^3,4^ Signs of glaucoma are detectable through detailed examination of optic nerve head (ONH) photographs, and automated image interpretation technologies, especially those based on artificial intelligence (AI), hold promise for enhancing reproducible, scalable screening frameworks.^5–8^ Therefore, developing robust methodologies to integrate and leverage AI into glaucoma screening protocols is critical to overcome the challenges of widespread implementation across diverse clinical environments.

Convolutional neural networks (CNNs) are a type of AI technology that have shown the capability to match or even exceed experts in interpretating fundus photographs to detect glaucoma, thereby creating an opportunity to improve the reproducibility and scalability of screening programs.^7,9,10^ Conventional CNN models often utilize supervised learning with transfer learning, enhancing their efficiency through pretraining on labeled non-ophthalmic images from the *ImageNet* database.^11,12^ In contrast, the novel RETFound foundation model, which is built on a vision transformer architecture (ViT), leverages self-supervised learning, being pretrained on 904,170 unlabeled fundus photographs and 1.4 million images from ImageNet, thereby developing a robust knowledge base adaptable to specific tasks via training on labeled images.^13^ RETFound has outperformed supervised learning-based ViTs in the classification of diabetic retinopathy, glaucoma, and exudative age-related macular degeneration, and demonstrated superior performance generalizability and efficacy with small training datasets.^13–17^ RETFound is also an effective feature extractor, capable of predicting cup-to-disc ratio (CDR) and circumpapillary retinal nerve fiber layer (RNFL) thickness from fundus photographs.^18^ Despite these results, guidelines for the optimal implementation of these AI models remain lacking as direct comparative studies on the effectiveness in glaucoma classification are limited.

In this study, we developed and tested RETFound and CNN models for classifying referable glaucoma based on ONH appearance using data from the Los Angeles County (LAC) Department of Health Services (DHS) teleretinal screening program.^19^ As the second largest municipal healthcare system in the United States, LAC DHS screens approximately 1,750 patients per month for select eye diseases, including diabetic retinopathy and glaucoma, through its teleretinal screening program.^20^ We evaluated the effects of image cropping and training set size and assessed the explainability and generalizability of the models–key challenges in developing AI models for screening environments. Through rigorous comparison of these architectures and development factors, our study seeks to provide insights into standardized methodologies for designing AI-assisted teleglaucoma screening programs, meeting a rapidly growing area of interest and need in modern eye care.

## Methods

This study was approved by the Institutional Review Boards of the University of Southern California. The study adhered to the tenets of the Declaration of Helsinki and complied with the Health Insurance Portability and Accountability Act.

### Data Collection

The LAC DHS administers a teleretinal screening program, serving approximately 1,750 incident diabetic patients a month at 17 hospital- and community-based sites.^19^ Participants are screened with dilated fundus photography, performed by trained photographers using the Topcon NW400 and NW8 (Topcon Corporation, Tokyo, Japan) and Canon CR-2 AF Digital (Canon U.S.A. Inc, Huntington, NY) cameras. Three standardized 45-degree photographs are taken: one centered on the optic nerve, one centered on the fovea, and one of the temporal macula.^19^ A team of 15 certified optometrists remotely review the photographs for diabetic retinopathy and referable glaucoma. Referable glaucoma is defined at a patient level for a cup-to-disc ratio (CDR) of 0.6 or greater. If pathology is detected, referrals are placed for in-person visits at LAC DHS eye clinics, where eyecare providers perform a complete evaluation for glaucoma.

All participants aged 18 years or older with at least one fundus photograph taken between January 4, 2016 and December 2, 2022 were eligible for this study. Photographs of the standardized field of view centered on the optic nerve were selected for inclusion. Photographs were excluded only if the optic nerve was not gradable due to complete obscuration by medial opacity or by not appearing in the field of the image. Photographs of low quality where the optic nerve was gradable were included. Photographs were labeled as demonstrating referable glaucoma or not according to the patient-level assessment made by the optometrists in the screening program. A second label was assigned to each photograph in the test dataset (i.e., eye-level) through independent grading by a panel of three fellowship-trained glaucoma specialists (V.N., B.W., B.Y.X.), adjudicated by majority rule (at least 2 of 3 for a consensus label). Data on demographics (age, ethnicity, race, gender) and medical history were extracted from medical records. Ethnic and racial identity was defined first by ethnicity (Hispanic or not), then the non-Hispanic group was divided by race (Black, White, Asian).

An external dataset was collected from routine clinic visits at the Roski Eye Institute of the University of Southern California. Photographs were taken using the ZEISS FF 450 and CLARUS (ZEISS, Oberkochen, Germany), Topcon TRC-NW8 (Topcon Corporation, Tokyo, Japan), and Optos California (Optos plc, Dunfermline, United Kingdon) cameras with a variety of fields of view. Participants included were aged 18 years or older with at least one fundus photograph taken between October 5, 2017 and July 30, 2023. Photographs were selected where the optic nerve was gradable and relatively centered. Cases of referable glaucoma were identified as participants examined by a fellowship-trained glaucoma specialist with a CDR of 0.6 or greater as documented in the electronic health record. Controls were identified as participants from any ophthalmology clinic with photographs reviewed by a fellowship-trained glaucoma specialist (K.B.) to confirm an intact neuroretinal rim with a CDR of less than 0.6.

All images and extracted data were curated in EyeAI, a data repository and computing platform built on the Deriva-ML framework.^21^ This allowed for reproducible data partitioning and algorithm development as described in the next section.

### Algorithm Development

The LAC DHS dataset was divided at the patient level into development (80%) and test (20%) datasets. The development dataset was further split for training (75%) and validation (25%). Some patients with multiple teleretinal screening visits were represented multiple times in the training and validation datasets, but reference labels by LAC DHS optometrists were unique for each visit. Multiple training subsets (N = 20, 400, 1000, 2000, 4000, or 6000 images) were created through random sampling of the training dataset to assess the effect of training set size on model performance. Stratified random sampling with equal allocation was used to ensure unbiased subset selection and to balance class distribution. The test dataset contained 1000 images from 500 patients with no overlap of patients with the development datasets. The external test dataset contained 300 images from 203 patients.

For algorithm development, labels of referable glaucoma were generalized to photographs of both eyes from the patient-level labels provided by the LAC DHS optometrists. All images were preprocessed by a proprietary software developed in Python, which has been described previously, and development datasets of cropped and uncropped images were created.^9^ After preprocessing, all images were reviewed manually to ensure appropriate cropping and centration.

Four deep learning models were developed on the cropped and uncropped datasets using two architectures. Two models were based on RETFound, a large ViT and foundation model, which has prior self-supervised learning on unlabeled images from ImageNet-1K and 904,170 color fundus photographs.^13^ The models were fine-tuned from the pretrained RETFound_mae_meh weights (available through https://github.com/rmaphoh/RETFound_MAE) to classify referable glaucoma with the default hyperparameters (i.e., 100 epochs, batch size of 16, base learning rate 5e-3, layer decay 0.65, etc.). Two additional models were based on the VGG-19 architecture, which was chosen for its high performance, efficiency with image-based data, and previous validation on the LAC DHS dataset.^9^ Specific details of the architectural adjustments and development specifications for the VGG-19 models are described in our prior publication.^9^ Separate models of both architectures were developed on the cropped and uncropped image datasets. Four models were also trained for each of the training subsets while holding constant the full validation dataset. During subset training, hyperparameters were not tuned and instead were held constant from models based on the full development dataset to reduce noise.

Each model was tested with cropped or uncropped images from the 1000-image test dataset, matching whether cropped or uncropped images were used in the training phase. Model performance was assessed using the refence labels by the panel of glaucoma specialists. Models trained on the full training dataset (N = 8,996) were also tested on the 300-image external test dataset, again using cropped or uncropped images matching the training phase.

50 LAC DHS images of referable glaucoma where predictions from the uncropped image RETFound and VGG-19 models disagreed were selected to assess the interpretability of each architecture and elucidate how they attended to different regions of the fundus photographs. xRAI saliency maps were generated for the predicted class using 10% and 25% saliency thresholds and the salient regions of the fundus photographs were reviewed by two fellowship-trained glaucoma specialists (K.B., B.Y.X.) to assess inclusion of relevant (i.e., the ONH and focal changes in RNFL reflectance) and irrelevant portions of the photographs.

### Statistical Analysis

Continuous measures were summarized by means and standard deviations, and categorical measures were summarized by proportions and percentages. Baseline demographic and clinical characteristics were compared between the development and test datasets using 2-tailed student t-tests or chi-square tests. Algorithm performance was compared using area under the receiver operating characteristic curve (AUC-ROC), area under the precision-recall curve (AUC-PR), F1 score, sensitivity, and specificity. DeLong’s method was used to generate confidence intervals and compare AUC-ROCs. Sensitivity and specificity were calculated at the optimal threshold as defined by Youden’s J statistic. Sensitivity was also calculated at a 95% specificity level to simulate the high specificity desired in a screening setting. Performance was assessed in demographic groups defined by race, ethnicity, gender, and age by comparing AUC-ROCs across these groups stratified in the test dataset. 60 years was selected as the group threshold for age as it was the nearest decade margin to the median age (59.2). Statistical tests with a *p*-value < 0.05 were considered significant. Statistical analyses were performed using R version 4.4.1 (The R Foundation).

## Results

The study included 12,998 images from 6,116 participants, which were split into a training dataset of 8,996 images, a validation dataset of 3,002 images, and a test dataset of 1,000 images. The mean age of participants in the development (combined training and validation) and test datasets were 56.8±10.5 and 57.3±10.3 years **(Table 1)**. There were more females than males (55.0% and 52.4% female, respectively). Hispanic was the majority ethnic group (Hispanic 68.1% and 70.6%, respectively). Demographic characteristics did not vary significantly between datasets (*p* ≥ 0.30). 37.1% and 50.0% of patients had referable glaucoma in the development and test datasets, respectively.

**Table 1.**
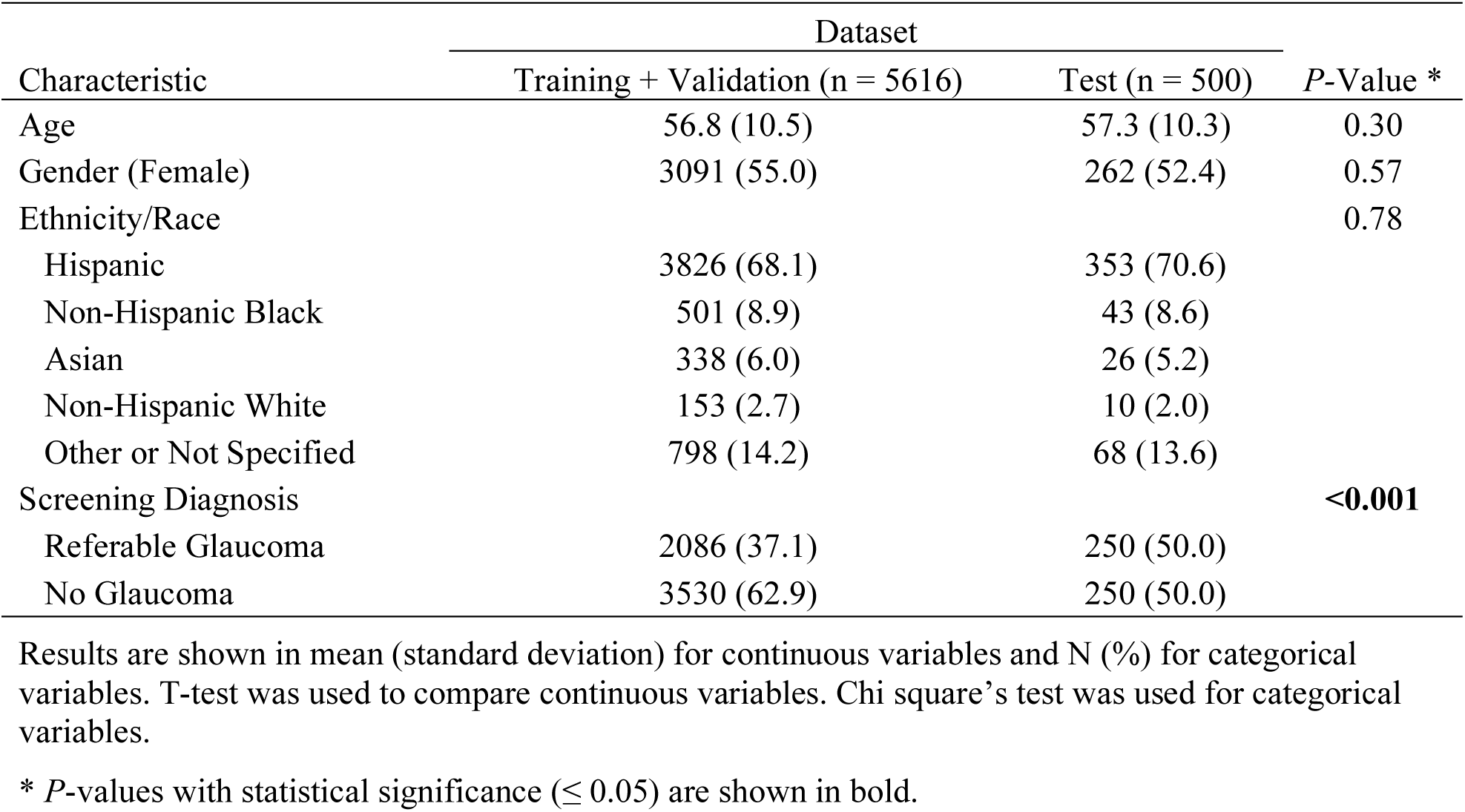
Demographic and Clinical Characteristics of the Study Population.

| Characteristic | Dataset |  | *P*-Value * |
| --- | --- | --- | --- |
|  | Training + Validation (n = 5616) | Test (n = 500) |  |
| Age | 56.8 (10.5) | 57.3 (10.3) | 0.30 |
| Gender (Female) | 3091 (55.0) | 262 (52.4) | 0.57 |
| Ethnicity/Race |  |  | 0.78 |
| Hispanic | 3826 (68.1) | 353 (70.6) |  |
| Non-Hispanic Black | 501 (8.9) | 43 (8.6) |  |
| Asian | 338 (6.0) | 26 (5.2) |  |
| Non-Hispanic White | 153 (2.7) | 10 (2.0) |  |
| Other or Not Specified | 798 (14.2) | 68 (13.6) |  |
| Screening Diagnosis |  |  | **<0.001** |
| Referable Glaucoma | 2086 (37.1) | 250 (50.0) |  |
| No Glaucoma | 3530 (62.9) | 250 (50.0) |  |

Results are shown in mean (standard deviation) for continuous variables and N (%) for categorical variables. T-test was used to compare continuous variables. Chi square's test was used for categorical variables.
\* *P*-values with statistical significance ( $\leq 0.05$ ) are shown in bold.

The VGG-19 architecture achieved higher threshold-independent performance than the RETFound architecture when trained using the full training set **(Figure 1**, **Table 2)**. The cropped image VGG-19 model reached an AUC-ROC of 0.924 (0.907, 0.940) and AUC-PR of 0.886, which was followed by the cropped image RETFound model (0.911 [0.892-0.930]; 0.848), the uncropped image VGG-19 model (0.898 [0.879-0.917]; 0.838), and the uncropped image RETFound model (0.889 [0.868-0.909]; 0.781). Pairwise comparisons of AUC-ROC demonstrated significant differences with cropped image models outperforming the uncropped image models of the same architectures (*p* ≤ 0.001) and the cropped image VGG-19 model outperforming the uncropped image RETFound model (*p* < 0.001). Pairwise comparisons of the cropped image RETFound and VGG-19 models (*p* = 0.07), cropped image RETFound and uncropped image VGG-19 models (*p* = 0.13), and uncropped image RETFound and VGG-19 models (*p* = 0.25) did not reach statistical significance. In contrast, the cropped image RETFound model had threshold-specific performance that exceeded the cropped image VGG-19 model **(Table 2)**. At their respective optimal thresholds, the cropped image RETFound model had a higher specificity than the cropped image VGG-19 model despite identical sensitivities (specificity 0.826 vs. 0.805 at sensitivity 0.882). It also had the highest F1 score among all models (0.801).

**Figure 1.**
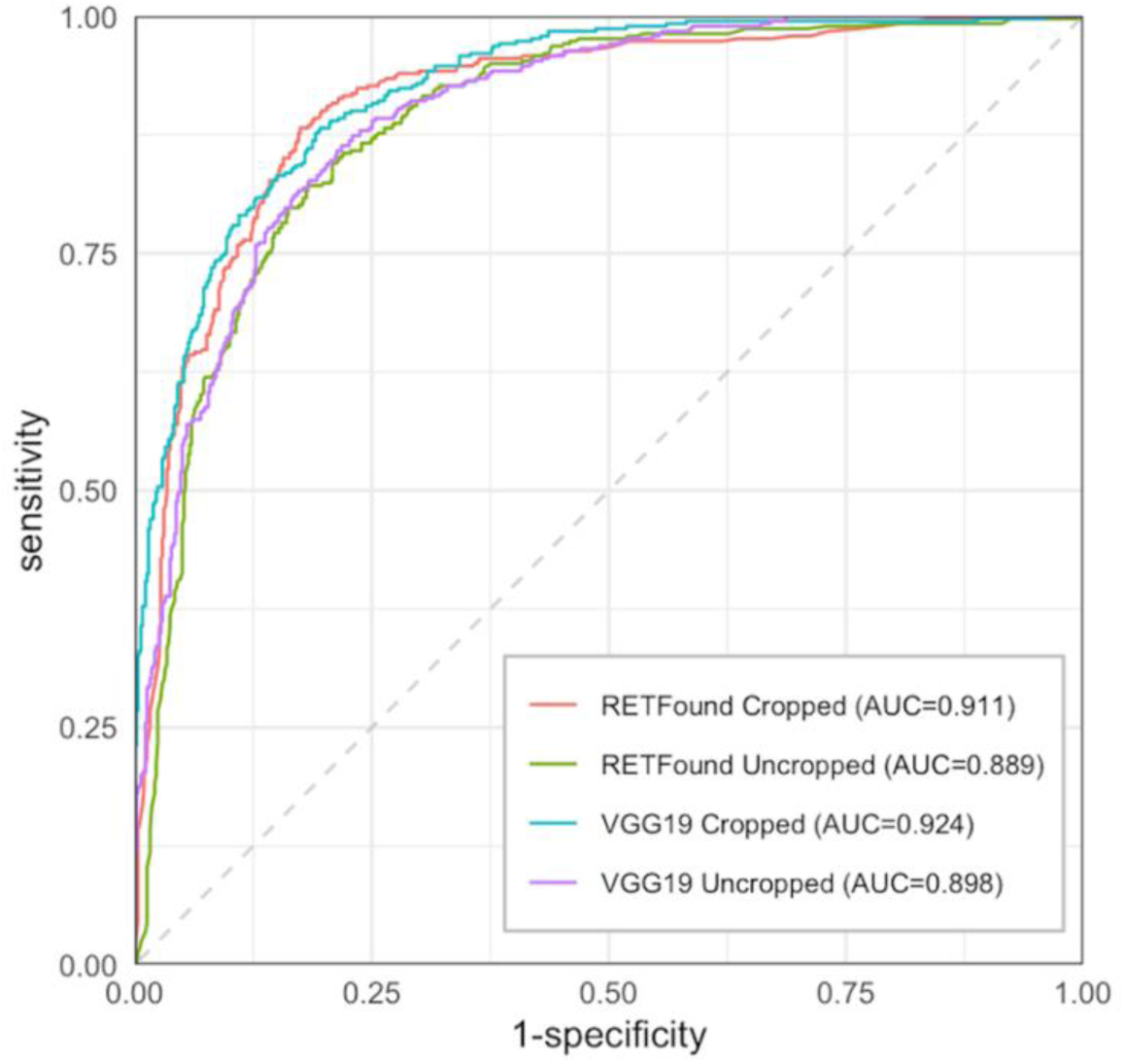
Receiver operating characteristic curves for RETFound and VGG-19 models against expert consensus labels.

**Table 2.** Performance Metrics of Models Tested on Expert Consensus Labels.

| Metric | Cropped Images |  | Uncropped Images |  |
| --- | --- | --- | --- | --- |
|  | RETFound | VGG-19 | RETFound | VGG-19 |
| AUC-ROC | 0.911 [0.892-0.930]* | 0.924 [0.907-0.940]<sup>†‡</sup> | 0.889 [0.868-0.909]*<sup>†</sup> | 0.898 [0.879-0.917]<sup>‡</sup> |
| AUC-PR | 0.848 | 0.886 | 0.781 | 0.838 |
| F1 Score | 0.801 | 0.757 | 0.769 | 0.759 |
| Specificity at Optimal Threshold | 0.826 [0.796-0.855] | 0.805 [0.774-0.835] | 0.817 [0.787-0.847] | 0.782 [0.748-0.813] |
| Sensitivity at Optimal Threshold | 0.882 [0.848-0.913] | 0.882 [0.848-0.913] | 0.822 [0.782-0.858] | 0.864 [0.829-0.895] |
| Sensitivity at 95% Specificity | 0.614 | 0.614 | 0.412 | 0.520 |

AUC: area under curve, PR: precision-recall curve, ROC: receiver operating characteristic curve
Results are shown with 95% confidence intervals when derivable. The DeLong method was used for AUC values. Pairwise comparisons AUC-ROC that had P-values of statistical significance ( $\leq 0.05$ ) are shown with matching superscripts (\*<sup>†‡</sup>). Stratified bootstrapping was used for sensitivity and specificity at the optimal threshold. The optimal threshold for ROCs was calculated using Youden's J statistic.

When models were developed on training datasets of varying size, a performance advantage in AUC-ROC was evident for the RETFound architecture over VGG-19 with 400 and 1000 training images **(Figure 2)**. Cropping increased VGG-19’s performance on these smaller training datasets (*p* < 0.03), whereas it did not benefit RETFound (*p* > 0.20). At 2000 and 4000 training images, performance converged for RETFound and VGG-19 (*p* > 0.07 for models paired by cropping status). Cropping benefitted RETFound on training datasets of 2000 images and larger (*p* < 0.03). The cropping benefit for VGG19 was present at all training dataset sizes (*p* < 0.05) except 2000 images (*p* = 0.58).

**Figure 2.**
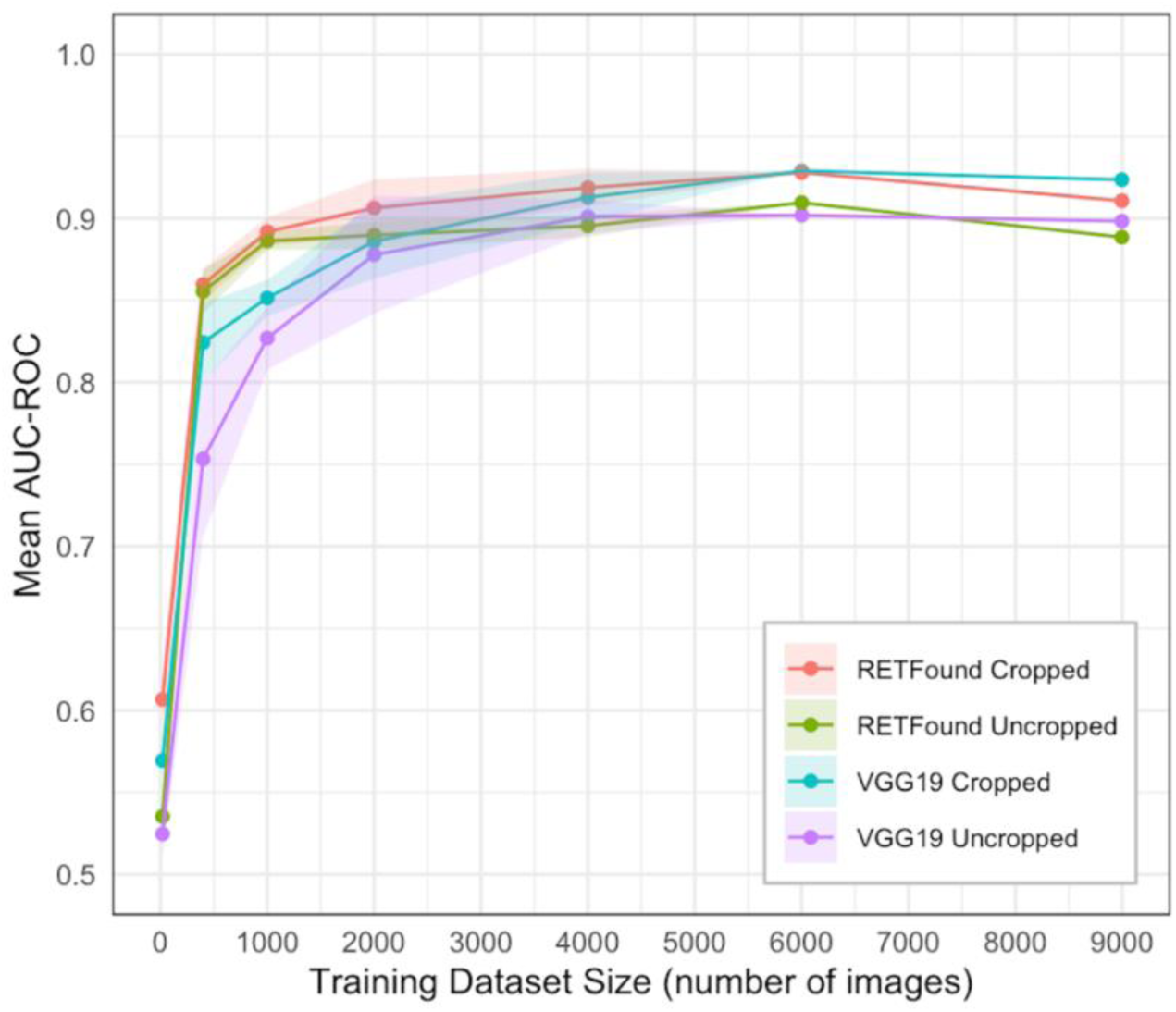
Mean AUC-ROCs for models of each architecture trained on increasing sizes of training datasets.

RETFound consistently performed better than VGG-19 on the external test dataset: the uncropped image RETFound model achieved an AUC-ROC of 0.886 (0.849, 0.924) and AUC-PR of 0.849, followed by the cropped image RETFound model (0.797 [0.746-0.848]; 0.761), the cropped image VGG-19 model (0.767 [0.712-0.822]; 0.813), and the uncropped image VGG-19 model (0.634 [0.571-0.697]; 0.641). The uncropped image RETFound model outperformed all other models in AUC-ROC (*p* < 0.001), had the best threshold-specific performance with the highest F1 score (0.812), and produced higher sensitivities at matched specificities, including a sensitivity of 0.707 when matched to the optimal specificity of the cropped image RETFound and VGG-19 models (0.880) **(Table 3, Supplemental Figure 1)**.

**Table 3.**
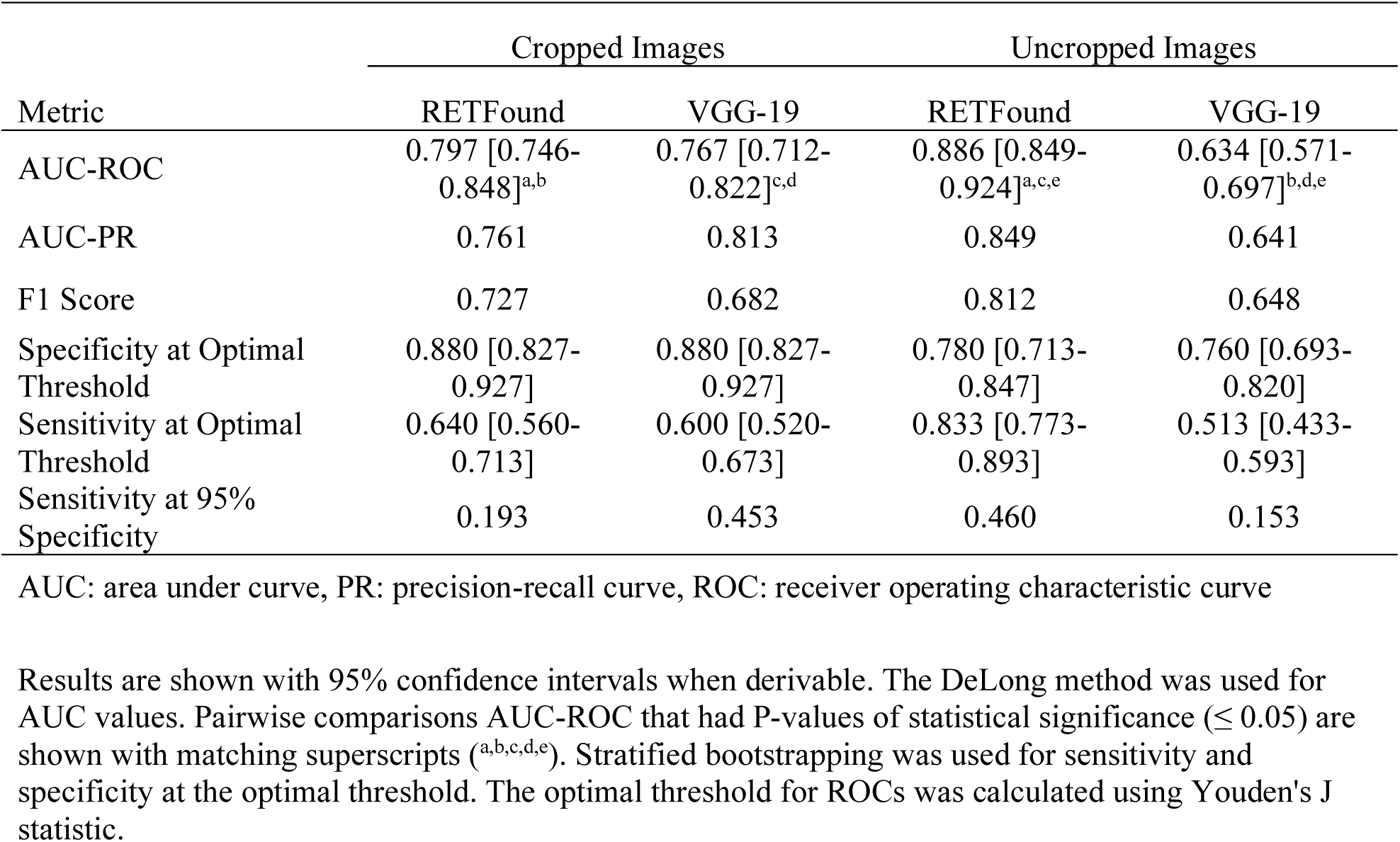
Performance Metrics of Models Tested on External Dataset.

| Metric | Cropped Images |  | Uncropped Images |  |
| --- | --- | --- | --- | --- |
|  | RETFound | VGG-19 | RETFound | VGG-19 |
| AUC-ROC | 0.797 [0.746-0.848]<sup>a,b</sup> | 0.767 [0.712-0.822]<sup>c,d</sup> | 0.886 [0.849-0.924]<sup>a,c,e</sup> | 0.634 [0.571-0.697]<sup>b,d,e</sup> |
| AUC-PR | 0.761 | 0.813 | 0.849 | 0.641 |
| F1 Score | 0.727 | 0.682 | 0.812 | 0.648 |
| Specificity at Optimal Threshold | 0.880 [0.827-0.927] | 0.880 [0.827-0.927] | 0.780 [0.713-0.847] | 0.760 [0.693-0.820] |
| Sensitivity at Optimal Threshold | 0.640 [0.560-0.713] | 0.600 [0.520-0.673] | 0.833 [0.773-0.893] | 0.513 [0.433-0.593] |
| Sensitivity at 95% Specificity | 0.193 | 0.453 | 0.460 | 0.153 |

AUC: area under curve, PR: precision-recall curve, ROC: receiver operating characteristic curve
Results are shown with 95% confidence intervals when derivable. The DeLong method was used for AUC values. Pairwise comparisons AUC-ROC that had P-values of statistical significance ( $\leq 0.05$ ) are shown with matching superscripts (<sup>a,b,c,d,e</sup>). Stratified bootstrapping was used for sensitivity and specificity at the optimal threshold. The optimal threshold for ROCs was calculated using Youden's J statistic.

When stratified by demographic group, the models demonstrated similar performance trends in AUC-ROC. All models, except the cropped RETFound model (*p* = 0.20), performed better on non-Hispanics than Hispanics (*p* < 0.04) **(Table 4)**. The models performed equally well in males and females (*p* > 0.28), and participants over and under the age of 60 years (*p* > 0.31).

**Table 4.** Performance by AUC-ROC Stratified by Demographic Group.

| Demographic Attribute | Group | n | Cropped Images |  | Uncropped Images |  |
| --- | --- | --- | --- | --- | --- | --- |
|  |  |  | RETFound | VGG-19 | RETFound | VGG-19 |
| Race | Asian | 52 | 0.939 [0.869, 1] | 0.953 [0.891, 1] | 0.908 [0.826, 0.989] | 0.866 [0.753, 0.978] |
|  | Non-Hispanic Black | 86 | 0.874 [0.792, 0.956]* | 0.922 [0.867, 0.976] | 0.871 [0.791, 0.952] | 0.919 [0.861, 0.977] |
|  | Non-Hispanic White | 20 | 0.980 [0.926, 1]*† | 0.961 [0.868, 1] | 0.961 [0.868, 1] | 0.941 [0.810, 1] |
|  | Hispanic | 706 | 0.902 [0.879, 0.926]† | 0.907 [0.885, 0.929]* | 0.874 [0.847, 0.900]* | 0.882 [0.857, 0.907]* |
|  | Other | 136 | 0.935 [0.888, 0.982] | 0.965 [0.939, 0.992]* | 0.930 [0.882, 0.978]* | 0.936 [0.898, 0.974]* |
| Ethnicity | Hispanic | 706 | 0.902 [0.879, 0.926] | 0.907 [0.885, 0.929]† | 0.874 [0.847, 0.900]† | 0.882 [0.857, 0.907]† |
|  | Non-Hispanic | 294 | 0.928 [0.897, 0.959] | 0.954 [0.933, 0.975]† | 0.919 [0.887, 0.952]† | 0.930 [0.903, 0.957]† |
| Gender | Female | 526 | 0.914 [0.888, 0.941] | 0.922 [0.899, 0.946] | 0.886 [0.857, 0.916] | 0.907 [0.881, 0.934] |
|  | Male | 448 | 0.904 [0.875, 0.933] | 0.918 [0.894, 0.943] | 0.888 [0.856, 0.920] | 0.886 [0.856, 0.915] |
| Age | > 60 years | 462 | 0.901 [0.871, 0.93] | 0.923 [0.900, 0.947] | 0.877 [0.845, 0.909] | 0.891 [0.862, 0.92] |
|  | ≤ 60 years | 538 | 0.920 [0.896, 0.944] | 0.923 [0.900, 0.946] | 0.898 [0.871, 0.925] | 0.904 [0.880, 0.929] |

AUC: area under curve, ROC: receiver operating characteristic curve
95% confidence intervals and *P*-values were derived using DeLong's method. Pairwise comparisons of sensitive groups within a single model that had significant *P*-values ( $\leq 0.05$ ) are shown with matching superscripts (\*†‡).

In the explainability analysis, all saliency maps (50 out of 50) for both architectures highlighted the optic nerve head at both saliency thresholds, regardless of the accuracy of the prediction **(Figure 3)**. All of the saliency maps (50 out of 50) also included small parts of the peripapillary region, connected to the optic nerve head, often following the contours and trajectories of vessels. The saliency maps for both models on two images included the fovea, but no clear foveal findings were evident to the human evaluators.

**Figure 3.**
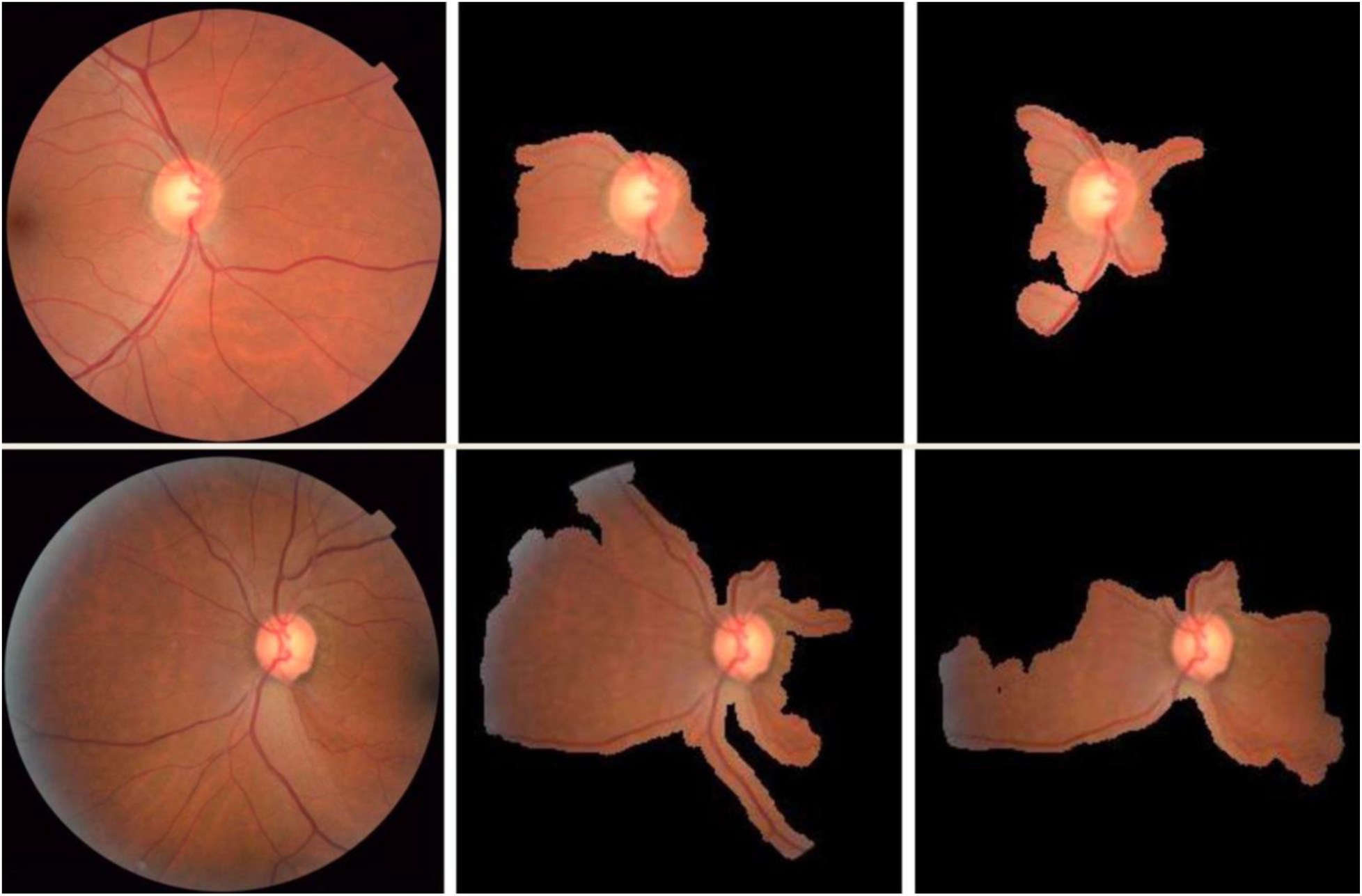
Example xRAI saliency maps at 10% (top) and 25% (bottom) thresholds: original (left), RETFound (center), and VGG19 (right).

## Discussion

Our study demonstrates that while conventional CNNs can detect referable glaucoma with performance comparable to RETFound, the latter’s extensive pretraining confers a significant advantage in scenarios with limited training data or domain variability. Specifically, RETFound outperforms CNNs when training datasets are small and when there is a shift in imaging conditions. We also found that cropping fundus images to focus on the optic nerve consistently enhanced classification accuracy across architectures, highlighting a straightforward yet critical step for optimizing AI-based screening. These findings provide novel insights into methodological standardization, addressing key knowledge gaps regarding AI deployment in real-world screening settings.

Our comparative analysis showed that RETFound and VGG-19 models have similar performance for detecting referable glaucoma in photographs from the LAC DHS teleretinal screening program. This parity in performance differs from two previous studies. Chuter et al., tested RETFound on cropped fundus photographs from the research setting, labeled according to ONH appearance of glaucoma, and found performance was better than a CNN (based on ResNet-50 architecture) previously reported by the same group, although the development sets differed between studies.^16^ Zhou et al. tested RETFound on publicly available glaucoma datasets, in which glaucoma was defined as manifest glaucoma based on clinical examination and visual field testing, and found performance was better than a model based on the same ViT architecture limited to pretraining on ImageNet.^13^ The higher AUC-ROCs and similar performance between architectures in our study may reflect that classification of referable glaucoma is easier than manifest glaucoma, since ground-truth labeling comes solely from the fundus photographs. Uniformity of the photograph acquisition protocol in the teleretinal screening program may have also helped VGG-19 achieve higher performance. These findings together suggest that the benefit of RETFound over other architectures varies by task and setting, which currently needs to be tested on a case-by-case basis.

Cropping fundus photographs to focus on the optic nerve demonstrated a consistent performance benefit across architectures in our study, which highlights its importance in the referable glaucoma classification task. However, the benefit of cropping has not been consistent across glaucoma classification studies. Zhen et al. reported that cropping modestly degraded CNN performance for glaucoma severity grading when applied to hospital-based fundus photographs, Saha et al. reported that cropping improved performance for binary glaucoma classification when applied to publicly available datasets, and Almeshrky et al. reported the effects of cropping varied depending on the CNN architecture and data source.^22–24^ Cropping has an inherent value of focusing the model on the most clinically relevant portion of the photograph—the optic nerve—although some of these studies and findings from Hemelings et al. suggest there is important information beyond the optic nerve region.^25^ Our explainability experiment demonstrated that, even in cases of disagreement, the RETFound and VGG-19 architectures both attended to the optic nerve and its immediate surroundings, suggesting that most salient information is at or close to the ONH. Another potential benefit to cropping is standardization of field of view, which may make CNN models more robust to external data. While our findings provide additional evidence in support of cropping, the optimal parameters and use cases for cropping warrant further systematic study.

The uncropped image RETFound model was the most robust to domain shift as evidenced by its significantly greater performance compared to other models on the external (USC) test dataset. We included the external test dataset to assess the effect of variations in camera type and image magnification and field of view. While the performance of the other models was degraded on the external compared to internal (LAC DHS) test dataset, performance of the uncropped image RETFound model remained stable in terms of AUC-ROC, AUC-PR, and F1 score. RETFound likely benefitted from the general robustness of the ViT backbone to distribution perturbation and shift and its pretraining with images from a variety of camera types.^13,26,27^ Our findings align with Zhang et al., who reported that RETFound exhibited the smallest performance drop for classifying diabetic retinopathy on external data.^15^ While cropping helped VGG-19, it adversely affected RETFound, suggesting that uncropped images allow better handling of camera type and image orientation variations. This insight enhances our understanding of architecture selection for dynamic real-world applications but also underscores the need to balance the robustness of foundation models with the standardization benefits conferred by cropping.

Foundation models inherently require less labeled training data, reducing barriers related to high-quality data curation for developing AI models. Although we observed improved label efficiency with RETFound, the advantage was less pronounced than reported in prior studies. Chuter et al. reported that RETFound outperformed a CNN trained on 9,473 images with a 2,000-image development set. In our study, both architectures trained on 2,000 images achieved high AUC-ROCs (RETFound median 0.907, VGG-19 median 0.887), with RETFound maintaining a statistically significant advantage (p = 0.01). With 4,000 images, VGG-19 matched RETFound’s performance, indicating that the benefit of foundation models is more prominent with smaller datasets. Our findings are consistent with those by Kuo et al., who found that the difference between RETFound and CNN performance in detecting diabetes-related OCT changes was most pronounced on small training datasets and converged at 500 images.^17^ Despite the prevailing belief that ViTs require more data due to fewer inductive biases, the foundation model architecture appears to mitigate this limitation, matching prior evidence and generally supporting the data-saving potential— although the magnitude of this advantage likely varies with task complexity.^13,16,27^

An additional benefit of foundation models is potential equity in performance across demographic groups, owing to training on extensive, diverse datasets. Pretraining RETFound on over 900,000 fundus images exposed it to a variety of retinal morphologies across multiple ethnicities (13.7% British, 14.9% Indian, 5.2% Caribbean, 3.9% African, 37.9% other, 24.4% unspecified). Our results showed consistent performance advantages for non-Hispanic populations, except for the cropped RETFound model, which demonstrated more balanced performance. Chen, et al. reported that the performance of RETFound in extracting latent features from uncropped images was better for non-Hispanic than Hispanic Whites.^18^ We consistently found similar worse performance among Hispanics compared to non-Hispanic Whites despite having a high representation of Hispanics in our training data for fine-tuning. The observed disparities across models may reflect morphological differences rather than bias in the pretraining dataset— highlighting the importance of considering biological variability in model design.

Our study has several limitations. First, the training dataset was labeled at the patient level for referable glaucoma, which may have reduced performance, particularly if images with CDR < 0.6 were labeled as referable based on the fellow eye. However, we previously demonstrated that our cropped VGG-19 model performs as well as if not better than ophthalmologists and optometrists despite this approach. Second, the predominantly Hispanic cohort limited our ability to assess race-specific effects, with smaller groups (e.g., Asians, Blacks, non-Hispanic Whites) exhibiting high performance variance. Third, although we included an external test dataset, labels were provided by the same glaucoma specialists as in the internal and external test sets, which limited our ability to evaluate label generalizability and subjective variability of CDR assessment across clinicians. Finally, incomplete glaucoma testing in our cohort restricts the reproducibility and objectivity of labels, though we used consensus expert ratings to mitigate subjective variability in evaluating optic nerve appearance.

In conclusion, both CNN and RETFound architectures demonstrate strong efficacy in classifying referable glaucoma, employing comparable classification strategies grounded in optic disc analysis. Foundation models like RETFound appear advantageous when training data are limited (<2000 images) and when domain shift is anticipated. While cropping enhances performance generally, it may diminish a model’s capacity to adapt to diverse imaging conditions, especially for foundation models optimized for robustness. These insights could inform best practices for designing AI screening tools, emphasizing the value of standardized preprocessing and architecture selection tailored to deployment environments. Moving forward, standardization of deployment methodologies, including image preprocessing, architecture selection, operating point validation and prospective testing, is essential to advance AI integration into clinical workflows effectively as has been done with other imaging modalities and ophthalmology use cases.^7,10,28,29^

## Data Availability

All data produced in the present study are available upon reasonable request to the authors.

## Acknowledgements

This work was supported by grant R01 EY035677 and K23 EY032985 from the National Eye Institute, National Institutes of Health, Bethesda, Maryland; a DHS-USC Safety Net Innovation Award from the Southern California Clinical and Translational Science Institute; a AI4Health Award from the University of Southern California; and an unrestricted grant to the Department of Ophthalmology from Research to Prevent Blindness, New York, NY.

---

**Supplemental Figure 1.**
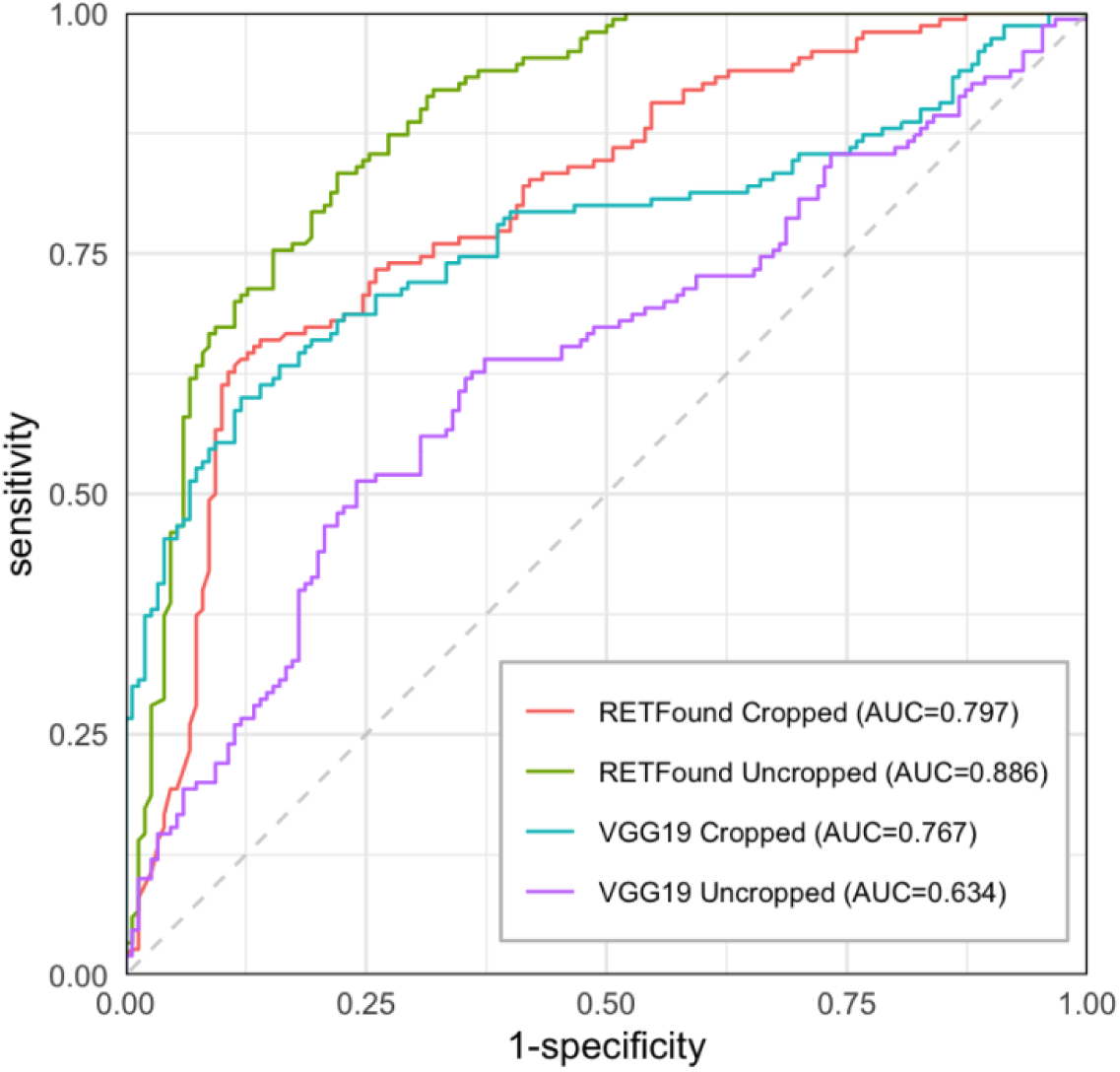
Receiver operating characteristic curves for RETFound and VGG-19 models on the external test dataset.

